# Naloxone Buyers Club: Overlooked Critical Public Health Infrastructure for Preventing Overdose Deaths

**DOI:** 10.1101/2021.11.14.21266221

**Authors:** Maya Doe-Simkins, Eliza Jane Wheeler, Mary C. Figgatt, T. Stephen Jones, Alice Bell, Peter J. Davidson, Nabarun Dasgupta

**Affiliations:** Remedy Alliance For the People, Oakland, California, USA; Gillings School of Global Public Health and Injury Prevention Research Center, University of North Carolina, Chapel Hill, NC USA; T. Stephen Jones Public Health Consulting, Florence, MA USA; Prevention Point Pittsburgh, Pittsburgh, PA USA; University of California, San Diego, CA USA

**Keywords:** opioid, overdose, naloxone, harm reduction, pharmaceutical, supply chain, standing orders

## Abstract

**Background:** Community-based naloxone distribution is an evidence-based pillar of overdose prevention. Since 2012, the naloxone Buyers Club facilitated purchase of low-cost naloxone by harm reduction and syringe service programs, the primary conduits for reaching people who use drugs. This innovative purchasing and mutual aid network has not been previously described.

**Methods:** We analyzed transactional records of naloxone orders (2017-2020, n=965), a survey of current Buyers Club members (2020, n=104), and mutual aid requests (2021, n=86).

**Results:** Between 2017 and 2020, annual orders for naloxone increased 2.6-fold. 114 unique harm reduction programs from 40 states placed orders for 3,714,110 vials of 0.4 mg/mL generic naloxone through the Buyers Club. States with most orders were: Arizona (600,000 vials), Illinois (576,800), Minnesota (347,450), California (317,200), North Carolina (315,040). Among programs that ordered naloxone in 2020, 52% (n=32) received no federal funding and ordered half as much as funded programs. During the 2021 shortage, mutual aid redistribution was common, with 80% participating as either a donor or recipient. Among 59 mutual aid requestors, 59% (n=35) were willing to accept expired naloxone; the clear preference was for generic injectable naloxone, 95% (n=56).

**Conclusions:** The naloxone Buyers Club is a critical element of overdose prevention infrastructure. Yet, barriers from corporate compliance officers and federal prescription-only status impede access. These barriers can be reduced by FDA removing the prescription requirement for naloxone and government funding for harm reduction programs.

## 1. INTRODUCTION

Opioid overdose has been a major driver of untimely mortality in the United States for two decades. Social isolation during the COVID-19 pandemic and fluctuations in unregulated drug supply have exacerbated the issue (Linas et al., 2021; Mason et al., 2021; Rodda et al., 2020; Slavova et al., 2020). In 2020, over 93,000 people lost their lives to overdose, about 70% from opioids, a 30% annualized increase (Ahmad et al., 2021). These deaths are largely preventable.

Providing the overdose antidote, naloxone (N-allylnoroxymorphone) is an evidence-based intervention (Davis and Carr, 2015; McDonald and Strang, 2016; Walley et al., 2013; Wheeler et al., 2015). People who use drugs are most likely to be on the scene when an overdose occurs. Naloxone kits and training include information on preventing, recognizing, and reversing overdoses, and how to access other health services. Naloxone distribution sites include syringe services programs, health departments, mobile outreach vans, churches, and other community locations (Freeman et al., 2018; Xu et al., 2018). Some programs receive federal or state funding, but no previous publication has documented the scope or impact of this support. We document this previously undescribed, critical, component of public health overdose prevention.

To improve naloxone access, the Opioid Safety and Naloxone Network (OSNN) formed in 2008. As of October 2021, the group had 1,082 members, including people who use drugs, legal and policy experts, harm reduction programs, advocates, public health officials, researchers, medical professionals, and government officials. OSNN is an unincorporated entity and receives no funding. Members of OSNN have been involved in developing advocacy and educational materials instrumental in increasing naloxone access.

A major barrier to naloxone access is price. Funding for harm reduction and syringe service programs, the primary distributors of naloxone to people who use drugs, has been limited and unstable (Green et al., 2012). The price of naloxone has increased substantially during the last decade, with wide variation between branded and generic formulations. The most affordable formulation is generic injectable single-dose 0.4 mg/mL in 1 mL vials (Gupta et al., 2016). It is administered intramuscularly, and packaged by programs with appropriate syringes, which are of different size than syringes used for intravenous drug use.

Styled after early AIDS antiretroviral purchasing groups (Berkowitz, 1999; DeChristoforo and Minor, 1997; James, 1996), the OSNN Buyers Club is a collective of more than a hundred organizations comprising the backbone of community-based naloxone distribution in the United States (Dasgupta et al., 2021). The primary purpose of the Buyers Club is to facilitate the purchase of naloxone. In 2012, led by the late Dan Bigg and three of the authors of this paper, we negotiated a favorable price for the lifesaving medication with Hospira (now part of Pfizer). For nearly a decade, the vast majority of community-based naloxone distribution has quietly relied on this arrangement (Schreiber, 2021). Programs place orders with the Buyers Club, which are batched weekly and sent to Pfizer. Pfizer ships the naloxone to the programs, who pay Pfizer directly.

The Buyers Club is an unfunded, volunteer effort, and does not impose any fees. In addition to placing the orders, the majority of staff time is spent on assisting programs in filling out the substantial paperwork required by the manufacturer, because naloxone is designated by US Food and Drug Administration (FDA) to be prescription-only.

At the request of Pfizer, since 2012, the Buyers Club has operated an access-controlled program. The negotiated agreement with Pfizer requires programs:

- Be a member of OSNN.
- Be a registered non-profit organization or have a fiscal sponsor.
- Distribute naloxone for free to people who use drugs, their friends and family, and direct service providers (e.g., harm reduction or syringe service programs)
- Refrain from re-selling naloxone or supplying institutions (hospitals, health care facilities, first responders).
- Sign a confidentiality agreement prohibiting price disclosure.
- Have a non-residential address to receive shipments.
- Have a prescriber provide a medical license number *and* Drug Enforcement Agency (DEA) number to sign off on the purchase and distribution of naloxone.

Those organizations able to meet these stringent criteria often support smaller organizations without medical staff. Buyers Club members with unrestricted funds often engage in mutual aid redistribution of naloxone to these smaller programs serving hard-to-reach and marginalized populations. In this paper, we document the role that mutual aid redistribution plays in ensuring continuity for smaller and innovative programs. A recent legal analysis clarified that federal law does not prohibit this practice (Davis, 2020).

This analysis was conducted in the context of a national shortage of naloxone. In April 2021, Pfizer notified the Buyers Club of a manufacturing issue at the lone factory manufacturing naloxone, unrelated to COVID vaccines (Kornfield, 2021). Other liquid injectables were also affected. Pfizer officially stated that the shortage would not be resolved until February 2022 (Pfizer Hospital, 2021). While institutional (e.g., hospital) purchasers can purchase other generic versions at higher prices, this is not viable for non-profit organizations. While some programs have been able to purchase more expensive injectable (retail $30 per kit, naloxone cost only) or intranasal formulations ($75 per kit) (Schreiber, 2021), this 15-to-30x higher price has exerted significant and sudden financial strain on small programs, grossly limiting the amount of the antidote distributed during the worst period of overdoses in the nation’s history (Amaral, 2021; Dalton, 2021; Snow, 2021; Godvin, 2021; Kornfield, 2021; Mckesson, 2021; Schreiber, 2021). As the shortage is ongoing, a full analysis of its impact is beyond scope. However, the shortage has cast light onto systematic failures in naloxone procurement and distribution.

This evaluation describes naloxone purchases, funding sources, mutual aid redistribution, and resource scarcity for community-based public health programs. While surveys of Buyers Club programs have been published previously (Wheeler et al., 2015), we use transactional and survey data from the Buyers Club for the first time to document this critical element of overdose prevention infrastructure.

## 2. MATERIALS AND METHODS

### 2.1 Data Sources

#### 2.1.1 Naloxone Ordering Data

From 2017, the Buyers Club maintained detailed electronic ordering logs for all participating programs (n=114). Longitudinal data included program name and location, dates of order and receipt, number of units, updated with post-order modifications and delivery exceptions.

#### 2.1.2 Survey Data

From January through March 2021, we surveyed Buyers Club programs to describe experiences with naloxone funding, purchases, and resource scarcity during 2020. All Buyers Club members active at the end of 2020 (n=104) were asked to participate in the survey. We asked all programs regardless of 2020 ordering about resource scarcity, mutual aid, and fundraising. We then asked programs that ordered naloxone in 2020 via the Buyers Club (n=71) about receipt of federal and state funding for naloxone, staff salary, and participant incentives in 2020, using structured questions and free text fields for additional details. The survey was conducted electronically, with data quality checks.

#### 2.1.3 Mutual Aid Data

We analyzed a separate administrative dataset to quantify naloxone requests (n=86) during the 2021 shortage, June 1 to October 31. The Buyers Club maintains a “Get Some/Got Some” electronic request form where members (and associated public health programs) running low on naloxone list the number of vials needed, and programs with extra supply offer willingness to share. These records are appended with supply resolutions by Buyers Club staff. The Open Society Foundations provided unrestricted funding to programs to encourage mutual aid redistribution. In addition to informing scarcity, mutual aid requests provided insight into willingness to use expired naloxone (Pruyn et al., 2019) and formulation preferences.

#### 2.1.4 Contextual Examples

Contextual examples are provided to highlight root causes of supply chain disruptions, exposing policy, legal, and regulatory barriers. Six examples from distinct levels of the supply chain, drawn from free text survey responses and operational experience, are briefly summarized.

### 2.2 Analytic Approach

We calculated descriptive statistics and constructed visualizations. Missing data are reported as item non-response. Medians and interquartile ranges (IQR) are reported due to mean-median inequality.(Zheng et al., 2017) Heatmaps of naloxone orders were constructed in Stata MP version 17 (College Station, Texas, USA) and visually inspected for patterns. Analytic code and output are available in a Jupyter Notebook (Supplementary Material) (Dasgupta, 2021).

### 2.3 Ethics and Participation

#### 2.3.1 Ethics Statement

This study was reviewed by the University of North Carolina Office of Human Research Ethics, which determined that it does not constitute human subjects research as defined under federal regulations (#21-2451).

#### 2.3.2 Participation of People with Lived Experience

People with lived experience of drug use, overdose reversal and naloxone distribution are represented across more than a hundred programs participating in the Buyers Club. Several authors have more than a decade of experience using naloxone and reversing overdoses.

## 3. RESULTS

### 3.1 Naloxone Ordering

Between 2017 and 2020, 114 unique harm reduction programs, from 40 states, placed 965 orders through the Buyers Club (Table 1) totaling 3,714,010 vials of 0.4 mg/mL naloxone in 1 mL vials. Orders placed via the Buyers Club (Figure 1) increased 2.6-fold from 506,120 vials in 2017, to 1,296,290 in 2020. While the average 4-year order volume was 76,151 vials (standard deviation: 136,118), the range between smallest and largest was five orders of magnitude, 20 vials to 600,000. The median order per program was 23,500 vials (IQR: 6,750 to 67,250); the mean-median inequality encourages us to report the latter going forward. The states with the most programs were: California (30), Washington (6), Michigan (6), Minnesota (5), Illinois (4) and North Carolina (4). The five states with programs ordering the most naloxone were: Arizona (600,000 vials), Illinois (576,800), Minnesota (347,450), California (317,200), and North Carolina (315,040).

**Table 1.**
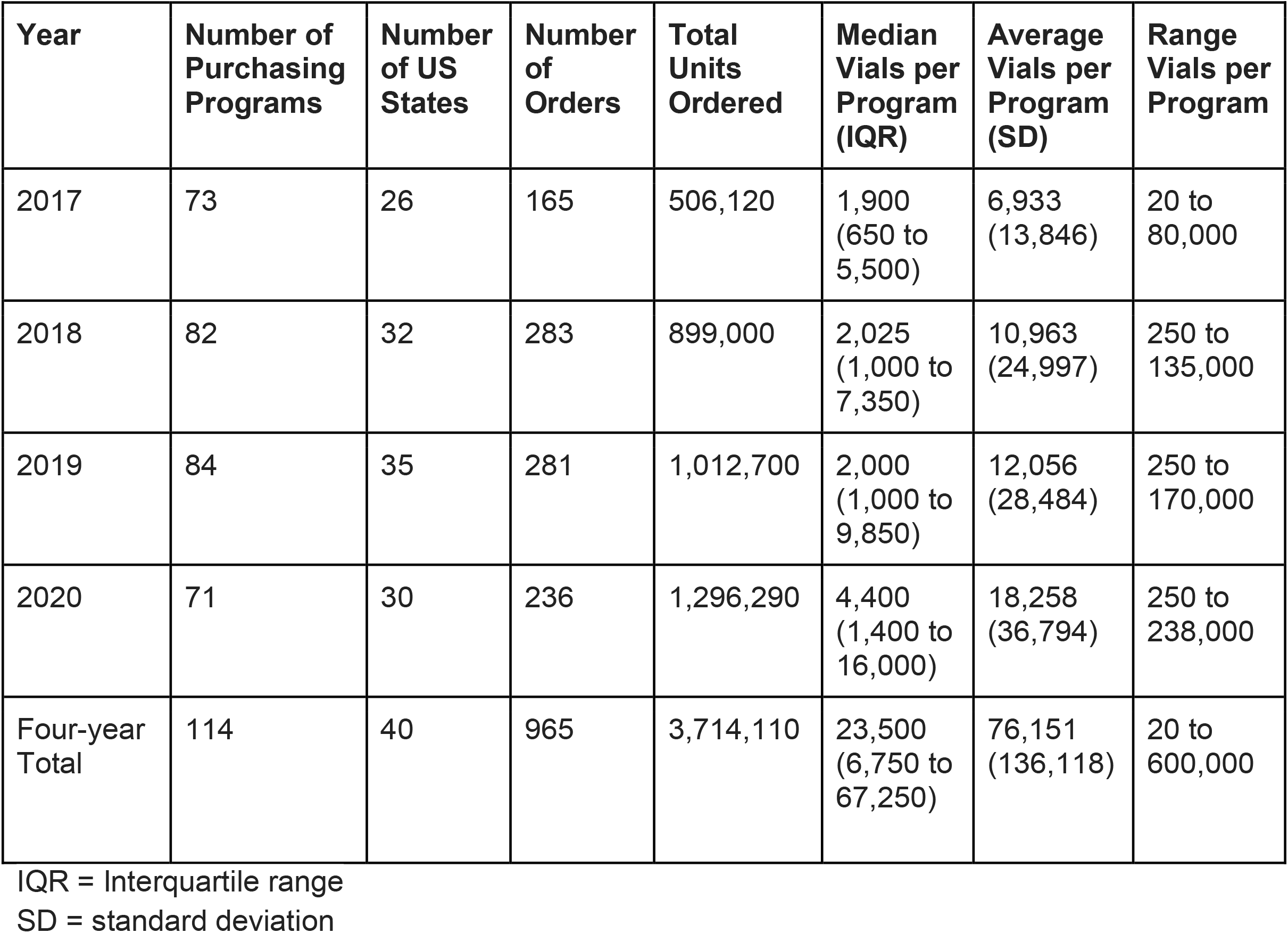
Annual Naloxone Orders by Harm Reduction Programs in the Buyers Club.

**Figure 1.**
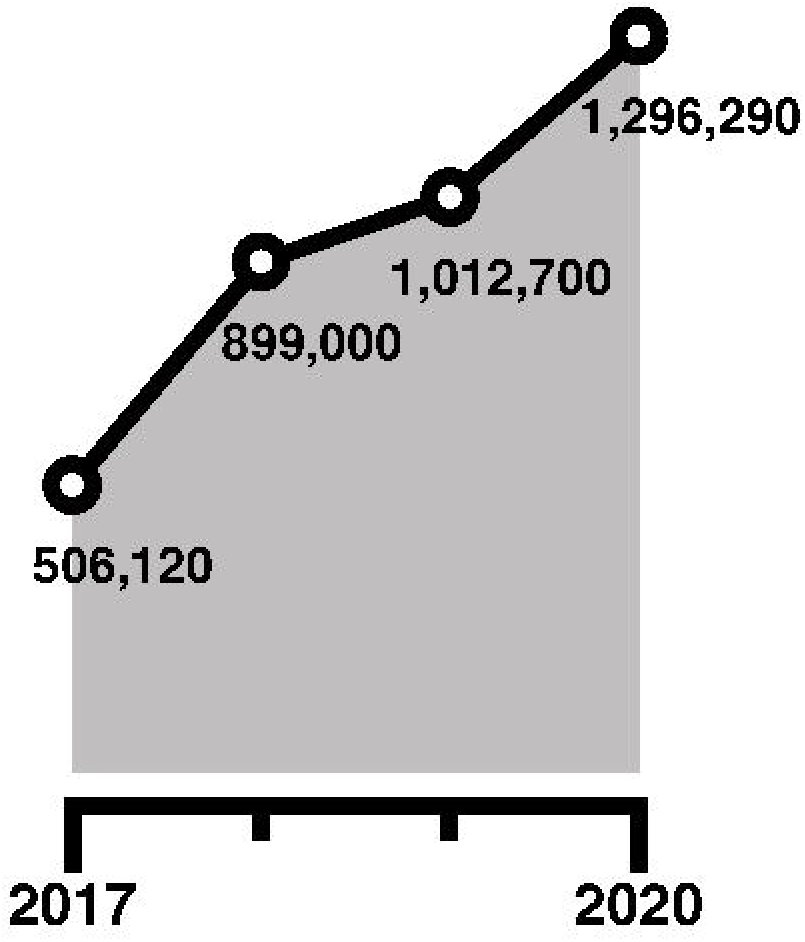
Annual naloxone orders by Buyers Club member programs. Yearly volume of orders by 114 harm reduction programs in the United States.

Visual inspection of longitudinal purchase histories in the heatmap (Figure 2) revealed heterogeneity across time (horizontal axis), over the 114 programs (vertical axis). Increasing color intensity with time represents greater demand moving from left to right. The whitespace also suggests that the Buyers Club supports many smaller programs with intermittent ordering. Many programs also receive naloxone through state health departments and corporate donations and use the Buyers Club as a backstop when other sources are insufficient. Larger programs make monthly purchases through the Buyers Club, reflecting limited storage capacity and locally high demand. April and December are peak months for ordering naloxone as calendar and fiscal year-limited funds expire. Some programs initially purchase monthly, and then choose to order larger amounts less frequently; this is depicted as lighter consecutive-month-runs followed by darker interval purchases. This may represent an initial vetting period as well as programmatic maturity, where storage space is increased, larger purchases become financially feasible, and growing demands of invoicing paperwork, all combine for less frequent but higher volume purchases.

**Figure 2.**
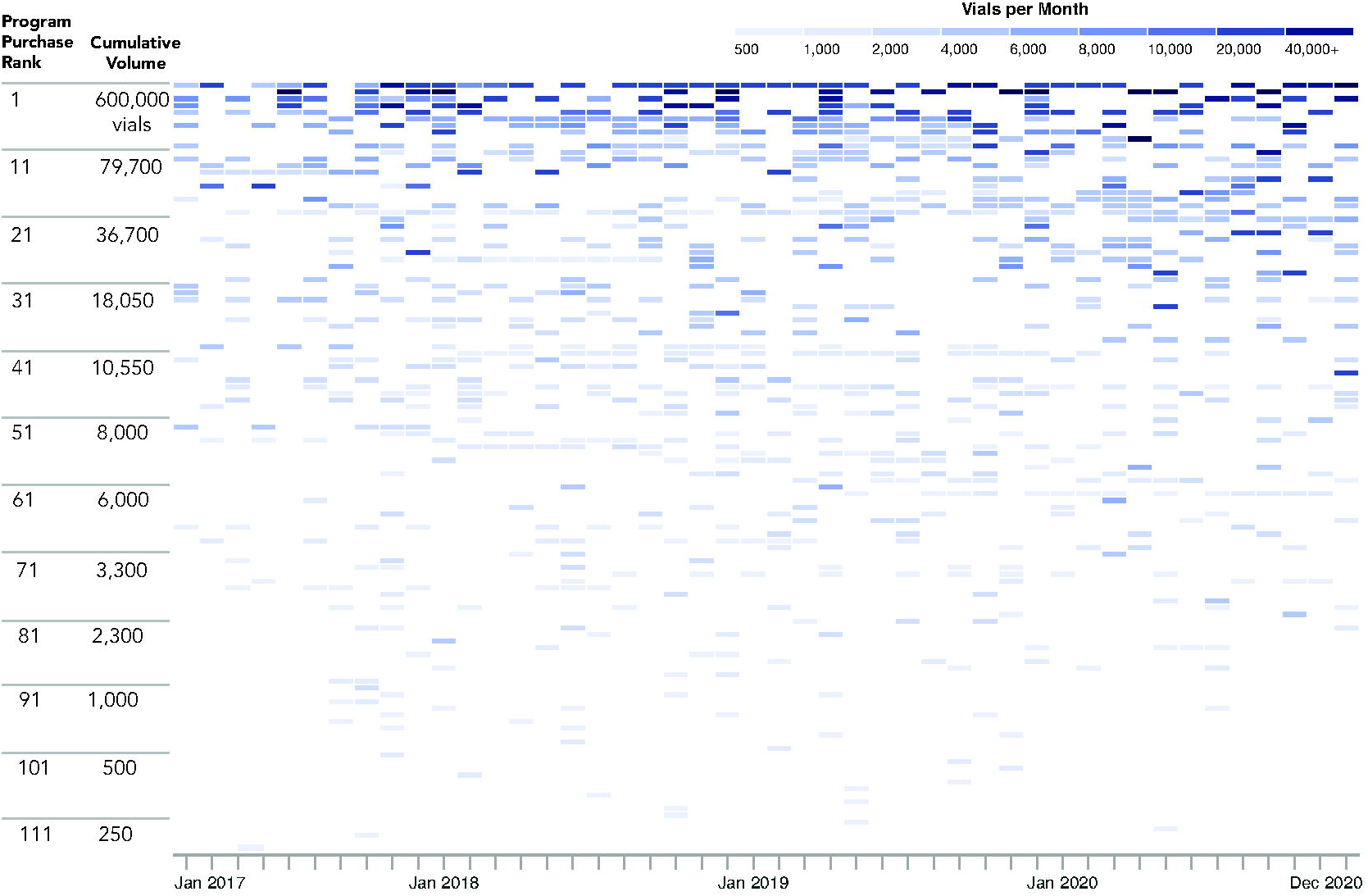
Heatmap of Longitudinal Naloxone Purchases by Buyers Club Member Programs. Each program’s (vertical axis) purchasing history was constructed as a longitudinal record (horizontal axis is time), with categorical colored indicators for monthly purchase volume. Each program is represented as a row, ranked in order of cumulative volume. Programs are ranked by descending cumulative order volume of naloxone purchased from 2017 to 2020, as designated on the left margin.

Programs tended to order a couple of times per year; the median was 6 separate orders (IQR: 2 to 12) during the 4-year study period, averaging 8.5 orders (standard deviation 7.6). During the study period, most programs (n=95, 83%) ordered more than once from the Buyers Club. The median time between orders was 62 days (IQR: 34 days to 119 days) among repeat purchasers.

### 3.2 Sources of Funding

At the end of 2020, there were 104 programs in the Buyer’s Club, operating in 40 states, all of which participated in the survey (response proportion 100%). A quarter of programs (25.3%, n=25) reported having to limit naloxone distribution due to affordable product scarcity. In 2020, one-in-five programs (20.0%, n=19) reported being required or encouraged to purchase more expensive formulations of naloxone with government grant funds.

The 71 programs that ordered naloxone through the Buyers Club in 2020 were asked about funding sources, with 67 responding (response proportion 94.4%). Among these 67 programs, only 48.4% (n=30) received any federal funding for naloxone (Table 2). The programs that received federal funds purchased more than twice the amount of naloxone (885,640 doses) as the programs that did not receive any federal funds (403,000 doses).

**Table 2.** Buyers Club Survey of Resources for Naloxone Distribution, 2020.

|  | Yes<br>N (%) | No<br>N (%) | Unknown,<br>Missing, or<br>Prefer not<br>to answer<br>N | Total |
| --- | --- | --- | --- | --- |
| *All Buyers Club Members (n=104)* |  |  |  |  |
| Did you ever need to limit naloxone distribution because you did not have enough naloxone or funds to purchase naloxone? | 25<br>(25.3%) | 74<br>(74.7%) | 5 | 104 |
| Were you asked to share, loan, gift your naloxone supply with another organization to meet their naloxone supply needs, OR, did you ask another organization to share, borrow, gift their supply with you to meet your naloxone supply needs? | 79<br>(79.8%) | 20<br>(20.2%) | 5 | 104 |
| Were you ever required/encouraged to use a portion of funds to purchase more expensive naloxone products? | 19<br>(20.0%) | 76<br>(80.0%) | 9 | 104 |
| *Buyers Club Members Recently Purchasing Naloxone (n=67\*)* |  |  |  |  |
| Did you get any federal funds to purchase naloxone?<br>(including federal funds passed through the state, county, or city) | 30<br>(48.4%) | 32<br>(51.6%) | 5 | 67 |
| Did you get any federal funds to cover staff, volunteer, participant time or incentives to distribute naloxone?<br>(including federal funds passed through the state, county, or city) | 29<br>(49.2%) | 30<br>(50.8%) | 8 | 67 |
| Did you get any state, county, or city government funds to purchase naloxone?<br>(excluding federal funds passed through the state, county, or city) | 29<br>(46.0%) | 34<br>(54.0%) | 4 | 67 |
| Did you get any state, county, city government funds to cover staff, volunteer, participant time or incentives to distribute naloxone?<br>(excluding federal funds passed through the state, county, or city) | 28<br>(45.2%) | 34<br>(54.8%) | 5 | 67 |
| Did you have to do fundraising to get enough money OR use unrestricted donations to purchase naloxone? | 30<br>(49.2%) | 31<br>(50.8%) | 6 | 67 |

\* Out of 71 programs ordering naloxone in 2020 in Table 1, four did not complete funding questions, resulting in a sample size of n=67.

Among programs who ordered naloxone via the Buyers Club in 2020 (n=67), half (49.2%, n=30) reported needing to do their own fundraising for purchasing naloxone (Table 2). Fundraising was accomplished through online crowdsourcing microfinance platforms and memorial pages for people who died of an overdose.

### 3.3 Mutual Aid Redistribution

From the funding survey, mutual aid redistribution was common practice in 2021, with 79.8% (n=79 out of 104) participating as either a donor or recipient of shared naloxone. During the period of Pfizer’s manufacturing-related shortage, June 1 to October 31, 2021, the Buyers Club received 59 requests for naloxone, and 27 offers to share naloxone. Offers to share were made without expectation for cost reimbursement, in compliance with Pfizer contracts. Buyers Club staff consolidated and coordinated the contribution and need requests, for example bundling offers to fill a larger request, or splitting contributions to fill smaller requests. Programs not in the Buyers Club were eligible to request donations, such as programs receiving secondary distribution from registered purchasers. Fifteen programs stated they needed money for shipping or storage. Through mutual aid redistribution 200,050 doses of naloxone were sent to programs in need.

### 3.4 Expired Naloxone

In an effort not to waste naloxone, recently expired naloxone has long been used by harm reduction programs to reverse overdoses successfully, and independent evaluations have confirmed shelf-life viability in excess of 5 years past printed expiration date (Lyon et al., 2006; Pruyn et al., 2019). However, some programs are prohibited from distributing expired naloxone by state or institutional rules. Among the 59 mutual aid requests for naloxone, 59% of programs (n=35) were willing to accept expired naloxone.

### 3.5 Formulation Preferences

The 59 mutual aid requests for naloxone in 2021 provide insight into which formulations are preferred by programs. Almost all programs (n=56, 95%) requested generic injectable naloxone. Narcan^®^ nasal spray was acceptable to 71% of programs (n=42), and only 37% (n=22) were willing to accept Evzio^®^ auto-injector (which was no longer marketed by the manufacturer at the time of the survey, and may have affected acceptability).

### 3.6 Contextual Experience

To provide context for quantitative findings, we provide six examples from Buyers Club members and associated public health programs, highlighting barriers to naloxone access at different levels of the supply chain. The prescription requirement limits naloxone access by community programs.

#### 3.6.1 Large Pharmaceutical Manufacturer

Pfizer requires DEA registration of the prescribing physician, even though naloxone is not a controlled substance. In their ordering system, one prescribing physician cannot provide their DEA authorization to multiple harm reduction programs, rendering statewide standing orders useless for naloxone procurement.

#### 3.6.2 Small Generic Manufacturer

In 2021, Hikma Pharmaceuticals’ generic naloxone was added to the Buyers Club product list at a discounted price. However, the company requires physicians to sign affidavits clarifying that the naloxone prescription explicitly authorizes “purchase,” and not solely “distribution.” This applies even to states with standing orders, some of which explicitly forbid purchasing.

#### 3.6.3 Pharmaceutical Distributor

When ordering from the pharmaceutical distributor, a convoluted fix was necessitated by the fact that harm reduction programs are ineligible for accounts in the McKesson division through which prescription drugs are sold, because they are not pharmacies. Therefore, naloxone has to be transferred between the distributor’s divisions (from prescription drug to Medical-Surgical), creating delays, supply chain vulnerabilities, and idiosyncratic order quotes or request denials from sales representatives.

#### 3.6.4 Regional Public Health Program

The Northwest Portland Area Indian Health Board serves 43 tribes in Washington, Oregon and Idaho. These rural and remote Native American communities have limited emergency medical care, and most have no access to in-person harm reduction services. While the organization can purchase naloxone, a legal assessment has prohibited them from mailing naloxone across state lines because the status is prescription-only.

#### 3.6.5 Local Public Health Programs

Public health initiatives in Connecticut and Michigan attempted to place vending machines for free naloxone in high overdose and service-scarce areas. Despite state standing orders, fear of enforcement of prescription drug fraud laws by FDA killed many target locations: Lawyers and organizational general counsels thwarted or stalled implementation because naloxone is prescription-only. Several vending machines continue to sit in a storage room, unused.

#### 3.6.6 Naloxone Donations

Even when cost is not an issue, regulatory and compliance issues leave overdose prevention efforts stymied. To receive a charitable donation of naloxone (Direct Relief, 2021), programs must comply with regulations for storing and dispensing medications, and have a licensed Medical Director or Pharmacist. In September 2021, the Buyers Club received a commitment from a pharmaceutical company of a 50,000 dose donation (Hikma Pharmaceuticals PLC, 2021). However, only *three* programs were able to complete the burdensome paperwork to receive the emergency donation.

## 4. DISCUSSION

Every 5 minutes someone in America dies of an overdose. For the first time, we document the critical public health infrastructure that supports community-based naloxone distribution. The Buyers Club facilitated purchase of over 3.7 million doses of naloxone over a four-year period. Despite this success, our data highlights substantial barriers to naloxone distribution, stemming from: 1) FDA classification of naloxone as “prescription-only”; and 2) inadequate financial investment in programs that reach marginalized people. Less than half of the Buyer’s Club programs received government funding for naloxone purchasing. Action by Congress and FDA to remove generic naloxone’s prescription requirement for harm reduction programs would prevent overdose deaths.

Our data reveal that the Buyers Club is anchored by a handful of very high volume programs, with a sizable middle tier of regular but moderate orderers, and a large tail of low volume intermittent purchasers. Whereas a commercial endeavor may deprioritize low volume purchasers, the Buyers Club administrators pay particular attention: These are specialized programs serving niche communities with resource scarcity. Some of these smaller programs serve remote Native American and Appalachian residents, Asian Americans, people recently leaving prison, rural church ministries, etc. These areas and populations already have markedly lower access to in-person harm reduction services, let alone health insurance to pay for naloxone in pharmacies. The Buyers Club plays a critical role in furthering equitable naloxone access that mainstream medicine and public health cannot accomplish.

As a result of naloxone scarcity, programs scrambled to implement a patchwork of various temporary strategies, including fundraising schemes, donations from other programs, or reducing naloxone distribution altogether. Government funding was inadequate and inconsistent. Despite the problems, Buyer’s Club organizations managed to increase naloxone access to vulnerable groups during the COVID-19 era, when many other health agencies suspended or reduced non-pandemic services. However, some programs faced dangerous scarcity. Secure funding access and naloxone supply will result in more life-saving naloxone in the hands of people who use drugs, the group most likely to use naloxone during an overdose.

State laws and executive orders intend for naloxone to be effectively treated as a non-prescription or over-the-counter product (Gertner et al., 2018; Lambdin et al., 2018; Sohn et al., 2019; Xu et al., 2018). Yet, it remains regulated as prescription-only at the federal level. This discrepancy (manifest as resistance from corporate and government compliance officers), greatly limits the ability of organizations to obtain and distribute naloxone and stifles innovation.

Government intervention is needed to provide a consistent, affordable stream of naloxone. Currently, government funding prioritizes naloxone for law enforcement, libraries, and schools. Compared to community-based programs, these constituencies do not reach populations in greatest need of naloxone as efficiently.

One success story is worth highlighting. The largest purchaser in the Buyers Club (600,000 doses) was Sonoran Prevention Works in Arizona. A recent analysis suggests that Arizona is the only state approaching saturation for adequate naloxone supply (Irvine et al., 2021). This program, supported by federal funding via the state health department, exclusively distributes cheap generic naloxone provided via the Buyers Club. More than 43,000 reversals have been recorded (Arizona Department of Health Services, 2021).

The Buyer’s Club is not the only source of community-distributed naloxone. The US Veterans Administration distributed naloxone to 285,279 patients from May 2014 to April 2021 (Oliva et al., 2021). Pfizer donated 1 million doses over four years, 2017 to 2021 (Direct Relief, 2021). The largest state government effort, in California, purchased 600,000 units from 2018 to 2021 on behalf of programs, funded by SAMHSA (California Department of Health Care Services, 2021). These purchases were insufficient: California programs purchased hundreds of thousands of additional doses through the Buyers Club. Cheap, generic naloxone via the Buyers Club is a backstop for inadequate government funding.

The majority of community-based naloxone distribution to people who use drugs is accomplished hand-to-hand, far from retail pharmacy shelves. While retail pharmacies are an intuitive venue for naloxone access, the observed volume of passed through this vast infrastructure is dwarfed by a comparatively smaller group of harm reduction programs. FDA and one branded manufacturer reported retail pharmacy sales of naloxone of 336,100 doses in 2017(Jiang, 2018) and 556,847 in 2018 (Guy et al., 2019), but was concentrated in a handful of states (US Food and Drug Administration, 2018).

During COVID, retail pharmacy sales of naloxone declined 26% (O’Donoghue et al., 2021), while demand from Buyers Club programs increased 29%. People who use drugs often do not feel comfortable purchasing naloxone in pharmacies (Antoniou et al., 2021). Pharmacists may label those asking for naloxone as “drug seeking”, further creating an unwelcome environment (Green et al., 2017). Experience from Australia suggests that only pharmacists already providing harm reduction services (e.g., syringes) are receptive to offering take-home naloxone (Nielsen and Olsen, 2021). Co-prescribing of naloxone with prescription opioids also faces low uptake (Haffajee et al., 2020). The main branded manufacturer (Emergent BioSolutions, Gaithersburg, Maryland) has forwarded a narrative emphasizing retail pharmacy as the primary avenue for over-the-counter naloxone for their expensive nasal spray (Knopf, 2020).

While offering the allure of rapid national distribution and a quick policy fix, international experience shows that the barriers to retail pharmacy distribution of naloxone are formidable (Donovan et al., 2019; Graves et al., 2019; Green et al., 2017; Olsen et al., 2019). On the other hand, research shows that people who use drugs are best-placed to reverse overdoses as they happen (Bennett et al., 2018). The US Veterans Administration reports 3.8% utilization rate for naloxone among law enforcement (Oliva et al., 2021); in one study, friends and family members of people who consume drugs, but are not drug users themselves, had a naloxone refill rate of only 1% (Bennett et al., 2018). In contrast, people who use drugs had a 11% return rate for refills after using naloxone to reverse an overdose (Bennett et al., 2018). Research from the United States (Wagner et al., 2014), Vietnam (Blackburn et al., 2017), and Norway (Eide et al., 2021) also shows that “Super-savers” naturally and consistently emerge during naloxone distribution. These empowered and motivated individuals report reversing multiple overdoses, having gained a positive reputation in the community. It is imperative that we evaluate and address different models of community distribution in order to get lifesaving medication into the right hands without exacerbating stigma.

As a retrospective review of transactional records and survey data, this study has limitations. Multivariable analysis was not undertaken because additional program characteristics were not available. An accurate market analysis is difficult because commercial data vendors do not capture naloxone distributed through all channels, such as donations. Some states with robust naloxone distribution programs (New Mexico, Massachusetts, New York) did not make any purchases from the Buyers Club, and are not represented. While mutual aid redistribution is common, some programs may be wary of disclosing these efforts because of fear of retribution from funding agencies. Naloxone distribution is not a panacea; not all overdoses are witnessed by someone able to administer the antidote (Stam et al., 2019). Despite these limitations, the data presented herein provide the first documentation of the naloxone Buyers Club, and can aid policy and regulatory decisions, as well as inform modelling for overdose prevention.

## 5. CONCLUSIONS

The naloxone Buyers Club is a unique, and previously unrecognized, facilitator of naloxone access. The network represents the backbone of community-based naloxone distribution in the United States. Yet, corporate compliance and legal barriers result in excessive administrative burdens that limit naloxone access. Reassessing funding priorities and removing the prescription requirement could reduce these barriers to ensure fairer and more efficient overdose prevention.

## Supporting information

Graphical abstract

COI disclosures

Contributorship statement

Funding

Highlights

programming code

## Data Availability

All data produced in the present work are contained in the manuscript.

https://doi.org/10.17615/tyd7-qy88

https://doi.org/10.17615/z548-vp45

## ACKNOWLEDGMENTS

We are grateful to the member organizations of the Buyers Club, and the people who informally redistribute naloxone to amplify saturation networks-usually via people who use drugs. We also thank the following people for comments on various portions and versions of the text of this manuscript: Corey Davis, Roxanne Saucier, Jessica Leston, and Daniel Raymond.

