## Supplementary material for "Naloxone Buyers Club: Overlooked Critical Public Health Infrastructure for Preventing Overdose Deaths": Graphical abstract

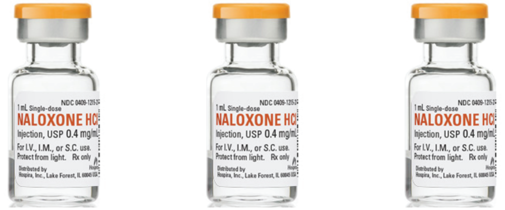

**Remedy Alliance // For The People**
**Estd. 2012**
RemedyAllianceFTP.org

### Data Sources

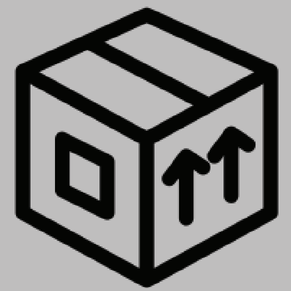

Naloxone Orders
N=965

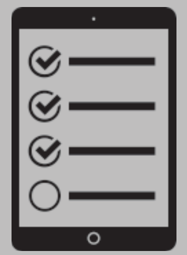

Member Survey
N=104
100% response

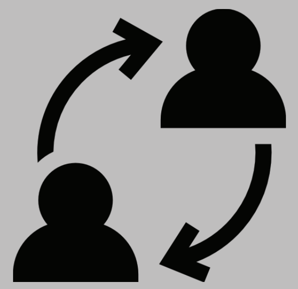

Mutual Aid Requests and Offers
N=86

**114 Buyers Club programs in 40 states ordered 3.7 million doses of generic injectable naloxone.**

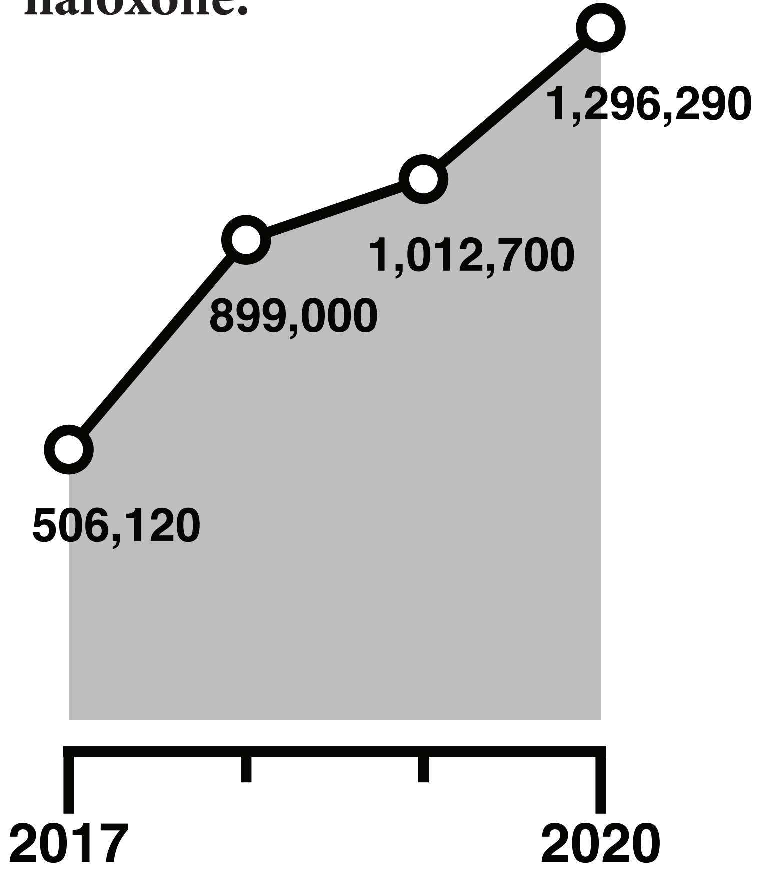


# 52%

**of harm reduction orgs received NO federal funding for naloxone and ordered 1/2 as much as programs with federal funding**

# 80%

**of programs shared naloxone through mutual aid**

### Barriers to Access

1) US FDA classification of naloxone as "prescription-only."

2) Corporate compliance fears of FDA enforcement.

3) Inadequate financial investment in programs that reach marginalized people.

4) Overemphasis in federal policy on retail pharmacy sales at the expense of harm reduction programs.
