## Supplementary material for "Naloxone Buyers Club: Overlooked Critical Public Health Infrastructure for Preventing Overdose Deaths": COI disclosures

**Conflict of Interest**

Dr. Dasgupta is a scientific advisor to the non-profit RADARS System of Denver Health and Hospitals Authority, a political subdivision of the State of Colorado (USA). Dr. Dasgupta does not accept personal compensation from any pharmaceutical company, distributor, or litigant. EJW, MD-S, PJD, AB, TSJ, MCF have no disclosures.
