## Supplementary material for "Naloxone Buyers Club: Overlooked Critical Public Health Infrastructure for Preventing Overdose Deaths": Contributorship statement

**Contributors**

EJW, MD-S, PJD, AB, TSJ conceptualized the study and developed the questionnaire. EJW and MD-S implemented data collection. ND, MD-S, and MCF conducted data analysis. All authors contributed to writing the manuscript.
