## Supplementary material for "Naloxone Buyers Club: Overlooked Critical Public Health Infrastructure for Preventing Overdose Deaths": Funding

**AUTHOR DISCLOSURES**

**Role of Funding Source**

Remedy Alliance (<https://remedyallianceftp.org/> - previously called OSNN Buyers Club) is an unfunded, non-profit entity. MD-S and EJW have volunteered their time to support the Buyers Club for a decade. No funding was received for preparation of this manuscript. Pfizer and Hikma have no direct financial relationship with the Buyers Club; all contracts are with individual programs. Neither company was aware of this manuscript until after submission.
