## Supplementary material for "Naloxone Buyers Club: Overlooked Critical Public Health Infrastructure for Preventing Overdose Deaths": Highlights

- Naloxone distribution through syringe programs is evidence-based prevention.
- The naloxone Buyers Club includes 114 harm reduction programs in 40 states.
- The Buyers Club facilitated purchases of 3.7 million doses between 2017 and 2020.
- Half members receive federal funding, necessitating mutual aid redistribution.
- To improve naloxone access, prescription-only designation should be reconsidered.
