## Supplementary material for "Naloxone Buyers Club: Overlooked Critical Public Health Infrastructure for Preventing Overdose Deaths": programming code

### Analysis of Naloxone Buyers Club Data

In [1]:
```
di "Stata MP"
version
di "Notebook generated on:$S_DATE at $S_TIME ET"
```

```
Stata MP

version 17.0

Notebook generated on: 2 Nov 2021 at 19:56:32 ET
```

Yellow highlight in PDF document denote numbers represented in the submited manuscript, including text, tables and figures.

#### Geographic Analysis by State

In [2]:
```
clear
import delimited "/Users/nabarun/Dropbox/Projects/Buyers Club manuscript/2017-2021 By State by year - B
y state by year.tsv", delimiter(tab) varnames(1)
rename v3 y2017
rename v4 y2018
rename v5 y2019
rename v6 y2020
foreach var of varlist y2017-y2020  {

        replace `var'=. if `var'==0

}

* data cleaning to remove notes
drop if state=="" | state=="YEAR TOTALS"

preserve
collapse (count) y2017-y2020
di "Number of states by year"
li
restore

foreach var of varlist y2017-y2020  {

        replace `var'=0 if `var'==.

}

gen total=y2017+y2018+y2019+y2020

gsort -total

di "Vials per state total for 4 years"
li state total
```

```
(encoding automatically selected: ISO-8859-1)
(10 vars, 54 obs)




(25 real changes made, 25 to missing)
(19 real changes made, 19 to missing)
(16 real changes made, 16 to missing)
(21 real changes made, 21 to missing)

(3 observations deleted)



Number of states by year


     +-------------------------------+
     | y2017   y2018   y2019   y2020 |
     |-------------------------------|
  1. |    26      32      35      30 |
     +-------------------------------+


(25 real changes made)
(19 real changes made)
(16 real changes made)
(21 real changes made)



Vials per state total for 4 years


     +-------------------------------+
     |                 state   total |
     |-------------------------------|
  1. |               Arizona  600000 |
  2. |              Illinois  576800 |
  3. |             Minnesota  347450 |
  4. |            California  317200 |
  5. |        North Carolina  315040 |
     |-------------------------------|
  6. |             Wisconsin  266700 |
  7. |                  Utah  196000 |
  8. |              Michigan  182900 |
  9. |               Indiana  165500 |
 10. |                 Maine  123650 |
     |-------------------------------|
 11. |            Washington  116600 |
 12. |          Rhode Island   58500 |
 13. |                Oregon   54400 |
 14. |                 Texas   51000 |
 15. |               Georgia   48400 |
     |-------------------------------|
 16. |              Colorado   48350 |
 17. |              Kentucky   36700 |
 18. |             Louisiana   27950 |
 19. |          Pennsylvania   27150 |
 20. |              Maryland   17700 |
     |-------------------------------|
 21. |                Nevada   15400 |
 22. | District of Columbia    14500 |
 23. |              Virginia   13850 |
 24. |                  Iowa   11100 |
 25. |         West Virginia   10500 |
     |-------------------------------|
 26. |        South Carolina   10250 |
 27. |              Missouri   10000 |
 28. |              New York    8150 |
 29. |         New Hampshire    6800 |
 30. |              Arkansas    6450 |
     |-------------------------------|
 31. |            New Jersey    6300 |
 32. |                  Ohio    6150 |
 33. |                Hawaii    5500 |
 34. |               Montana    2700 |
 35. |                 Idaho    2500 |
     |-------------------------------|
 36. |              Oklahoma    2300 |
 37. |          North Dakota    1800 |
 38. |             Tennessee     800 |
 39. |           Connecticut     800 |
 40. |               Florida     250 |
     |-------------------------------|
 41. |           Mississippi       0 |
 42. |                Kansas       0 |
 43. |               Alabama       0 |
 44. |                Alaska       0 |
 45. |              Delaware       0 |
     |-------------------------------|
 46. |               Vermont       0 |
 47. |              Nebraska       0 |
 48. |          South Dakota       0 |
 49. |         Massachusetts       0 |
 50. |            New Mexico       0 |
     |-------------------------------|
 51. |               Wyoming       0 |
     +-------------------------------+
```

#### Table 1

### 2017

In [3]:
```
/// 2017
clear
import delimited "/Users/nabarun/Dropbox/Projects/Buyers Club manuscript/OSNN Master 2021 - OSNN 2017 O
rders.tsv"

format customernumber %15.0g

drop if customernumber==.

* Distinct Programs
distinct programname

* Number of US States
distinct state

preserve
        collapse (sum) vials
        tab vials
restore

preserve
        collapse (sum) vials, by(programname)
        summ vials, detail
restore

* One observation from March 2017 is missing in the total dataset (Reno, NV)
* Create single observation dataset to append later
keep if vials==20
save missing2017, replace
```

```
(encoding automatically selected: ISO-8859-1)
(10 vars, 186 obs)


(21 observations deleted)


             |        Observations
             |      total   distinct
-------------+----------------------
 programname |        165         73


       |        Observations
       |      total   distinct
-------+----------------------
 state |        165         26



(sum) vials |      Freq.     Percent        Cum.
------------+-----------------------------------
     506120 |          1      100.00      100.00
------------+-----------------------------------
      Total |          1      100.00




                         (sum) vials
-------------------------------------------------------------
      Percentiles      Smallest
 1%           20             20
 5%          100             50
10%          250            100       Obs                  73
25%          650            100       Sum of wgt.          73

50%         1900                      Mean           6933.151
                        Largest       Std. dev.      13846.32
75%         5500          42000
90%        15000          43000       Variance       1.92e+08
95%        42000          57000       Skewness       3.331729
99%        80000          80000       Kurtosis       14.90401


(164 observations deleted)

file missing2017.dta saved
```

### 2018

In [4]:
```
clear
import delimited "/Users/nabarun/Dropbox/Projects/Buyers Club manuscript/OSNN Master 2021 - TOTAL 2018
 Orders.tsv", varnames(1)

drop if programname==""

* Fix program names
        replace programname="Baltimore Harm Reduction Coalition (BHRC)" if programname=="Baltimore Harm
 Reduction Coalition (formerly Baltimore Student HRC)"
        replace programname="PONI" if regexm(programname,"PONI")
        replace programname="End Overdose" if regexm(programname,"End Overdose")
        replace programname="Face to Face" if regexm(programname,"Face to Face")
        replace programname="Univ of Utah" if regexm(programname,"Utah")

* Distinct Programs Addresses
distinct programname

* Number of US States
distinct state


* Count vials
destring numberofvialsnotboxesorcasesinmu,  g(vials) force ignore(",")
        order vials, a(numberofvialsnotboxesorcasesinmu)

preserve
        collapse (sum) vials
        tab vials
restore

preserve
        collapse (sum) vials, by(programname)
        summ vials, detail
restore
```

```
(encoding automatically selected: ISO-8859-1)
(11 vars, 286 obs)

(3 observations deleted)

(3 real changes made)

(3 real changes made)

(1 real change made)

(4 real changes made)

(6 real changes made)


             |        Observations
             |      total   distinct
-------------+----------------------
 programname |        283         82


       |        Observations
       |      total   distinct
-------+----------------------
 state |        283         32

numberofvialsnotboxesorcasesinmu: character , removed; vials generated as long



(sum) vials |      Freq.     Percent        Cum.
------------+-----------------------------------
     899000 |          1      100.00      100.00
------------+-----------------------------------
      Total |          1      100.00




                         (sum) vials
-------------------------------------------------------------
      Percentiles      Smallest
 1%          250            250
 5%          250            250
10%          300            250       Obs                  82
25%         1000            250       Sum of wgt.          82

50%         2025                      Mean           10963.41
                        Largest       Std. dev.      24997.23
75%         7350          80000
90%        27500          85000       Variance       6.25e+08
95%        60000         130000       Skewness       3.579118
99%       135000         135000       Kurtosis       16.01498
```

### 2019

In [5]:
```
clear
import delimited "/Users/nabarun/Dropbox/Projects/Buyers Club manuscript/OSNN Master 2021 - Total 2019
 Orders.tsv", delimiter(tab) varnames(1)

drop if programname==""
* Distinct Programs Addresses
distinct programname


* Count vials
rename numberofvialsnotboxesorcasesinmu vials

preserve
        collapse (sum) vials
        tab vials
restore

preserve
        collapse (sum) vials, by(programname)
        summ vials, detail
restore
```

```
(encoding automatically selected: ISO-8859-1)
(8 vars, 285 obs)

(4 observations deleted)


             |        Observations
             |      total   distinct
-------------+----------------------
 programname |        281         84




(sum) vials |      Freq.     Percent        Cum.
------------+-----------------------------------
    1012700 |          1      100.00      100.00
------------+-----------------------------------
      Total |          1      100.00




                         (sum) vials
-------------------------------------------------------------
      Percentiles      Smallest
 1%          250            250
 5%          300            250
10%          300            250       Obs                  84
25%         1000            250       Sum of wgt.          84

50%         2000                      Mean           12055.95
                        Largest       Std. dev.      28484.43
75%         9850          65000
90%        36800          67500       Variance       8.11e+08
95%        63000         162000       Skewness       4.120229
99%       170000         170000       Kurtosis        21.4545
```

### 2020

In [6]:
```
clear
import delimited "/Users/nabarun/Dropbox/Projects/Buyers Club manuscript/OSNN Master 2021 - Total 2020
 Orders.tsv", delimiter(tab) varnames(1)


drop if programname==""
* Distinct Programs Addresses
distinct programname


* Count vials
rename numberofvialsnotboxesorcasesinmu vials

preserve
        collapse (sum) vials
        tab vials
restore

preserve
        collapse (sum) vials, by(programname state)
        summ vials, detail
restore
```

```
(encoding automatically selected: ISO-8859-1)
(9 vars, 238 obs)

(2 observations deleted)


             |        Observations
             |      total   distinct
-------------+----------------------
 programname |        236         71




(sum) vials |      Freq.     Percent        Cum.
------------+-----------------------------------
    1296290 |          1      100.00      100.00
------------+-----------------------------------
      Total |          1      100.00




                         (sum) vials
-------------------------------------------------------------
      Percentiles      Smallest
 1%          250            250
 5%          400            250
10%          700            400       Obs                  71
25%         1400            400       Sum of wgt.          71

50%         4400                      Mean           18257.61
                        Largest       Std. dev.      36794.04
75%        16000          90000
90%        39640         104000       Variance       1.35e+09
95%        90000         150000       Skewness       4.012288
99%       238000         238000       Kurtosis       21.52023
```

#### Full dataset analysis

Includes data processing for figure

##### All year data for last row

In [7]:
```
clear all
import delimited "/Users/nabarun/Dropbox/Projects/Buyers Club manuscript/2017-2021 By State by year - W
orksheet.tsv", delimiter(tab) varnames(1)
drop if year==.
destring vials,  replace force ignore(",")
drop if year==2021

* Add in missing observation
append using missing2017

drop v6-v9 naloxone month

gen date2=date(date,"MDY")
format date2 %td
order date2, a(date)
gen month = month(date2)

di "total vials"
preserve
        collapse (sum) vials
        tab vials
restore
```

```
(encoding automatically selected: ISO-8859-1)
(10 vars, 1,013 obs)

(2 observations deleted)

vials: character , removed; replaced as long

(47 observations deleted)

(variable v9 was str3, now str8 to accommodate using data's values)


(1 missing value generated)


(1 missing value generated)

total vials



(sum) vials |      Freq.     Percent        Cum.
------------+-----------------------------------
    3714110 |          1      100.00      100.00
------------+-----------------------------------
      Total |          1      100.00
```

#### Data cleaning for Figure 2

In [8]:
```
clear all
import delimited "/Users/nabarun/Dropbox/Projects/Buyers Club manuscript/2017-2021 By State by year - W
orksheet.tsv", delimiter(tab) varnames(1)
drop if year==.
destring vials,  replace force ignore(",")
drop if year==2021

* Add in missing observation
append using missing2017

drop v6-v9 naloxone month

gen date2=date(date,"MDY")
format date2 %td
order date2, a(date)
gen month = month(date2)


gen monthseq = month(date2) if year(date2)==2017
        replace monthseq = month(date2)+12 if year(date2)==2018
        replace monthseq = month(date2)+24 if year(date2)==2019
        replace monthseq = month(date2)+36 if year(date2)==2020
                order monthseq, a(date2)

preserve
        collapse (sum) vials, by(monthseq)
restore

bysort programname: egen total = total(vials)

sort date2
bysort programname: gen orders=_n
```

```
(encoding automatically selected: ISO-8859-1)
(10 vars, 1,013 obs)

(2 observations deleted)

vials: character , removed; replaced as long

(47 observations deleted)

(variable v9 was str3, now str8 to accommodate using data's values)


(1 missing value generated)


(1 missing value generated)

(801 missing values generated)

(283 real changes made)

(281 real changes made)

(236 real changes made)
```

In [9]:
```
di "Generate ranks for Figure 2"
preserve
        collapse (max) total, by(programname)
                gsort -total
                        gen group=_n
                list total
                                drop total
                                        save grouprank, replace
restore
```

```
Generate ranks for Figure 2




     +--------+
     |  total |
     |--------|
  1. | 600000 |
  2. | 535000 |
  3. | 259500 |
  4. | 240440 |
  5. | 196000 |
     |--------|
  6. | 160450 |
  7. | 153000 |
  8. | 120500 |
  9. | 114450 |
 10. | 114000 |
     |--------|
 11. |  79700 |
 12. |  67250 |
 13. |  58500 |
 14. |  50000 |
 15. |  49400 |
     |--------|
 16. |  48400 |
 17. |  45000 |
 18. |  40600 |
 19. |  37000 |
 20. |  36750 |
     |--------|
 21. |  36700 |
 22. |  35200 |
 23. |  35000 |
 24. |  30300 |
 25. |  27850 |
     |--------|
 26. |  27600 |
 27. |  27450 |
 28. |  26500 |
 29. |  25000 |
 30. |  23500 |
     |--------|
 31. |  18050 |
 32. |  17850 |
 33. |  16400 |
 34. |  15200 |
 35. |  14500 |
     |--------|
 36. |  14200 |
 37. |  14100 |
 38. |  13550 |
 39. |  13000 |
 40. |  11200 |
     |--------|
 41. |  10550 |
 42. |  10100 |
 43. |  10000 |
 44. |  10000 |
 45. |  10000 |
     |--------|
 46. |   9900 |
 47. |   9250 |
 48. |   9000 |
 49. |   8900 |
 50. |   8150 |
     |--------|
 51. |   8000 |
 52. |   7000 |
 53. |   6900 |
 54. |   6750 |
 55. |   6600 |
     |--------|
 56. |   6450 |
 57. |   6300 |
 58. |   6300 |
 59. |   6300 |
 60. |   6050 |
     |--------|
 61. |   6000 |
 62. |   5750 |
 63. |   5500 |
 64. |   5400 |
 65. |   4650 |
     |--------|
 66. |   4500 |
 67. |   4200 |
 68. |   4000 |
 69. |   3900 |
 70. |   3800 |
     |--------|
 71. |   3300 |
 72. |   2900 |
 73. |   2850 |
 74. |   2700 |
 75. |   2700 |
     |--------|
 76. |   2650 |
 77. |   2500 |
 78. |   2500 |
 79. |   2450 |
 80. |   2300 |
     |--------|
--more--
```

In [10]:
```
merge m:1 programname using grouprank, nogen
```

```
    Result                      Number of obs
    -----------------------------------------
    Not matched                             0
    Matched                               965
    -----------------------------------------
```

In [11]:
```
describe
```

```
Contains data
 Observations:           965
    Variables:            16
--------------------------------------------------------------------------------
Variable      Storage   Display    Value
    name         type    format    label      Variable label
--------------------------------------------------------------------------------
date            str10   %10s                  Date
date2           float   %td
monthseq        float   %9.0g
year            int     %8.0g                 Year
programname     str75   %75s                  Program Name
state           str2    %9s                   State
vials           long    %10.0g                Vials
customernumber  double  %15.0g                Customer number
address         str49   %49s                  Address
city            str22   %22s                  City
zip             str21   %21s                  Zip
v10             long    %12.0g
month           float   %9.0g
total           float   %9.0g
orders          float   %9.0g
group           float   %9.0g
--------------------------------------------------------------------------------
Sorted by:
     Note: Dataset has changed since last saved.
```

#### Programs per State

In [12]:
```
preserve
        keep if orders==1
    gen counter=1
        collapse (sum) counter (max) total, by(state)
        gsort -counter
        li counter state
restore
```

```
(851 observations deleted)


     +-----------------+
     | counter   state |
     |-----------------|
  1. |      30      CA |
  2. |       6      MI |
  3. |       6      WA |
  4. |       5      MN |
  5. |       4      NC |
     |-----------------|
  6. |       4      IL |
  7. |       3      MD |
  8. |       3      WV |
  9. |       3      ME |
 10. |       3      WI |
     |-----------------|
 11. |       3      NV |
 12. |       3      PA |
 13. |       3      OR |
 14. |       2      IN |
 15. |       2      SC |
     |-----------------|
 16. |       2      TX |
 17. |       2      NH |
 18. |       2      CO |
 19. |       2      LA |
 20. |       2      IA |
     |-----------------|
 21. |       2      TN |
 22. |       2      ND |
 23. |       2      VA |
 24. |       2      OH |
 25. |       1      AR |
     |-----------------|
 26. |       1      ID |
 27. |       1      FL |
 28. |       1      NY |
 29. |       1      RI |
 30. |       1      DC |
     |-----------------|
 31. |       1      HI |
 32. |       1      MO |
 33. |       1      CT |
 34. |       1      AZ |
 35. |       1      GA |
     |-----------------|
 36. |       1      KY |
 37. |       1      NJ |
 38. |       1      UT |
 39. |       1      MT |
 40. |       1      OK |
     +-----------------+
```

#### Table 1 - Last Row

In [13]:
```
di "Unique programs"
distinct programname
```

```
Unique programs


             |        Observations
             |      total   distinct
-------------+----------------------
 programname |        965        114
```

In [14]:
```
distinct state
```

```
       |        Observations
       |      total   distinct
-------+----------------------
 state |        965         40
```

#### Reordering of Vials by programs

In [15]:
```
preserve
        collapse (max) orders, by(group)
        tab orders
    di "Number of Orders"
        summ orders, detail
restore
```

```
      (max) |
     orders |      Freq.     Percent        Cum.
------------+-----------------------------------
          1 |         19       16.67       16.67
          2 |         10        8.77       25.44
          3 |         10        8.77       34.21
          4 |          7        6.14       40.35
          5 |          7        6.14       46.49
          6 |          6        5.26       51.75
          7 |          3        2.63       54.39
          8 |          4        3.51       57.89
          9 |          7        6.14       64.04
         10 |          5        4.39       68.42
         11 |          2        1.75       70.18
         12 |          7        6.14       76.32
         13 |          3        2.63       78.95
         14 |          3        2.63       81.58
         15 |          3        2.63       84.21
         16 |          4        3.51       87.72
         17 |          1        0.88       88.60
         18 |          1        0.88       89.47
         19 |          1        0.88       90.35
         21 |          4        3.51       93.86
         22 |          2        1.75       95.61
         23 |          1        0.88       96.49
         28 |          1        0.88       97.37
         30 |          1        0.88       98.25
         34 |          1        0.88       99.12
         38 |          1        0.88      100.00
------------+-----------------------------------
      Total |        114      100.00

Number of Orders


                        (max) orders
-------------------------------------------------------------
      Percentiles      Smallest
 1%            1              1
 5%            1              1
10%            1              1       Obs                 114
25%            2              1       Sum of wgt.         114

50%            6                      Mean           8.464912
                        Largest       Std. dev.      7.593873
75%           12             28
90%           19             30       Variance        57.6669
95%           22             34       Skewness       1.397875
99%           34             38       Kurtosis       5.090944
```

#### Table 1 - Last row for all programs

In [16]:
```
di "Vials per Program across all years"
summ total, detail
```

```
Vials per Program across all years


                            total
-------------------------------------------------------------
      Percentiles      Smallest
 1%          350             20
 5%         1750            100
10%         2700            250       Obs                 965
25%         6750            250       Sum of wgt.         965

50%        23500                      Mean           76151.01
                        Largest       Std. dev.      136117.7
75%        67250         600000
90%       196000         600000       Variance       1.85e+10
95%       535000         600000       Skewness       2.842785
99%       600000         600000       Kurtosis       10.69448
```

#### Figure 1: Orders per year

In [17]:
```
preserve
    collapse (sum) vials, by(year)
    format vials %12.0f
    li
    collapse (sum) vials
    li
restore
```

```
     +----------------+
     | year     vials |
     |----------------|
  1. | 2017    506100 |
  2. | 2018    899000 |
  3. | 2019   1012700 |
  4. | 2020   1296290 |
  5. |    .        20 |
     +----------------+


     +---------+
     |   vials |
     |---------|
  1. | 3714110 |
     +---------+
```

#### Figure 2

Code below but won't generate the chart image in Jupyter.

In [18]:
```
#delimit ;
 label define yrmth
      1 "Jan 2017"   13 "Jan 2018"
      25 "Jan 2019"  37 "Jan 2020"
   , replace ;
 #delimit cr

label values monthseq yrmth


quietly: summarize monthseq
 return list
 local count = `r(max)' + 1

 local xlab
    forval i=1/`count'{
        local xlab "`xlab' `=`i'-0.5' `" "`:lab (monthseq) `i''" "'"
    }

 #delimit ;
 heatplot vials i.group i.monthseq,

   ytitle("")
   xtitle("")

    cuts(@min 500 1000 2000 4000 6000 8000 10000 20000 40000 @max)

   legend(subtitle("{bf}Vials per Order          ",
        span
        size(1.75)
       )
     pos(1)
     ring(1)
     rows(1)
     keygap(0.5)
     colgap(0)
     size(1.5)
     symysize(0.85)
     symxsize(7)
     order(1 "500" 2 "1,000" 3 "2,000" 4 "4,000" 5 "6,000" 6 "8,000" 7 "10,000" 8 "20,000" 9 "40,000+")

     stack
    )


   xlabel(`xlab',
     nogrid
     labsize(1.75)
     labcolor(gs5)
    )
xscale(extend)
yscale(
     noline
     alt
     reverse
    )

   ylabel(,
     angle(horizontal)
     labgap(-145)
     labsize(2)
     noticks
     labcolor(gs5)
     nogrid
    )

   graphregion(margin(l=22 r=2))
   plotregion(margin(b=0 t=0))

   p(
    lcolor(white)
    lwidth(0.1)
    lalign(center)
   )


 colors(sfso, reverse)
 scheme(plotplain)

;

 #delimit cr
```

```
scalars:
                  r(N) =  964
              r(sum_w) =  964
               r(mean) =  26.00829875518672
                r(Var) =  160.9241262824076
                 r(sd) =  12.68558734479439
                r(min) =  1
                r(max) =  48
                r(sum) =  25072
```

#### Table 2: Survey Results

In [19]:
```
import delimited "/Users/nabarun/Dropbox/Projects/Buyers Club manuscript/OSNN Master 2021 - 2020 survey
 results.tsv", delimiter(tab) varnames(1) clear

** Remove erroneous data entry after close of survey
gen date2=date(timestamp,"MDY hms")
format date2 %td
order date2, a(timestamp)
drop if date2>date("30mar2021","DMY")

describe
```

```
(encoding automatically selected: ISO-8859-1)
(27 vars, 115 obs)



(11 observations deleted)


Contains data
 Observations:           104
    Variables:            28
-------------------------------------------------------------------------------
Variable      Storage   Display    Value
    name         type    format    label      Variable label
-------------------------------------------------------------------------------
timestamp       str18   %18s                  Timestamp
date2           float   %td
in2020didyoug~o str10   %10s                  In 2020, did you get any federal
                                                funds to purchase naloxone
                                                (include federal fun
howmuchofyour~s str20   %20s                  How much of your entire 2020
                                                naloxone supply did the federal
                                                funds (including fe
v4              str10   %10s                  In 2020, did you get any federal
                                                funds to cover
                                                staff/volunteer/participant time
howmuchofthes~t str14   %14s                  How much of the salary/incentives
                                                did the federal funds (including
                                                federal funds
in2020didyoug~t str10   %10s                  In 2020, did you get any
                                                state/county/city government
                                                funds to purchase naloxone
v7              str10   %10s                  In 2020, did you get any
                                                state/county/city government
                                                funds to cover staff/volun
fundingmixfor~e str31   %31s                  Funding mix for naloxone
fundingmixfor~s str31   %31s                  Funding mix for salary/incentives
in2020didyouh~t str10   %10s                  In 2020, did you have to do
                                                fundraising to get enough money
                                                OR use unrestricted
in2020didyoue~x str3    %9s                   In 2020, did you ever need to
                                                limit naloxone distribution
                                                because you did not ha
in2020wereyou~f str3    %9s                   In 2020, were you asked to
                                                share/loan/gift your naloxone
                                                supply with another org
wereyoueverre~u str10   %10s                  Were you ever required/encouraged
                                                to use a portion of funds to
                                                purchase more exp
isthereanythi~t strL    %9s                   Is there anything else you would
                                                like to say about overdose
                                                prevention funding?
programname     str77   %77s                  Program name
didyoumakeana~u str10   %10s                  Did you make a naloxone purchase
                                                through this buying club in
                                                2020?
whydidyounotm~s strL    %9s                   Why did you not make a naloxone
                                                purchase through this buying
                                                club in 2020? (Chec
ifyoudontknow~a str42   %42s                  If you don't know if the funds are
                                                federal but you DO know the name
                                                of the fundi
purchases       long    %12.0g                2017 purchases
v20             long    %12.0g                2018 purchases
v21             long    %12.0g                2019 purchases
v22             long    %12.0g                2020 purchases
total1720purc~s long    %12.0g                Total 17-20 purchases
change1718      float   %9.0g                 % change 17-18
change1819      float   %9.0g                 % change 18-19
change1920      float   %9.0g                 % change 19-20
changeoldestm~r str7    %9s                   % change oldest-most recent year
-------------------------------------------------------------------------------
Sorted by:
     Note: Dataset has changed since last saved.
```

"Did you ever need to limit naloxone distribution because you did not have enough naloxone/funds to purchase naloxone?"

In [20]:
```
tab in2020didyoueverneedtolimitnalox, m sort
```

```
   In 2020, |
    did you |
  ever need |
   to limit |
   naloxone |
distributio |
  n because |
you did not |
         ha |      Freq.     Percent        Cum.
------------+-----------------------------------
         No |         74       71.15       71.15
        Yes |         25       24.04       95.19
            |          5        4.81      100.00
------------+-----------------------------------
      Total |        104      100.00
```

"Were you asked to share/loan/gift your naloxone supply with another organization to meet their naloxone supply needs OR did you ask another organization to share/borrow/gift their supply with you to meet your naloxone supply needs?"

In [21]:
```
tab in2020wereyouaskedtoshareloangif, m sort
```

```
   In 2020, |
   were you |
   asked to |
share/loan/ |
  gift your |
   naloxone |
supply with |
another org |      Freq.     Percent        Cum.
------------+-----------------------------------
        Yes |         79       75.96       75.96
         No |         20       19.23       95.19
            |          5        4.81      100.00
------------+-----------------------------------
      Total |        104      100.00
```

"Were you ever required/encouraged to use a portion of funds to purchase more expensive naloxone products?"

In [22]:
```
tab wereyoueverrequiredencouragedtou, m sort
```

```
   Were you |
       ever |
required/en |
 couraged to |
      use a |
 portion of |
   funds to |
   purchase |
   more exp |      Freq.     Percent        Cum.
------------+-----------------------------------
         No |         76       73.08       73.08
        Yes |         19       18.27       91.35
            |          6        5.77       97.12
 Don't know |          3        2.88      100.00
------------+-----------------------------------
      Total |        104      100.00
```

#### Table 2: Limit to Programs that Purchased Naloxone in 2020

In [23]:
```
tab didyoumakeanaloxonepurchasethrou
keep if didyoumakeanaloxonepurchasethrou=="Yes"
```

```
    Did you |
     make a |
   naloxone |
   purchase |
    through |
this buying |
    club in |
      2020? |      Freq.     Percent        Cum.
------------+-----------------------------------
         No |         37       35.58       35.58
        Yes |         67       64.42      100.00
------------+-----------------------------------
      Total |        104      100.00

(37 observations deleted)
```

"Did you get any federal funds to purchase naloxone (include federal funds passed through the state/ county/city)?"

In [24]:
```
tab in2020didyougetanyfederalfundsto, m sort
```

```
   In 2020, |
did you get |
any federal |
   funds to |
   purchase |
   naloxone |
   (include |
federal fun |      Freq.     Percent        Cum.
------------+-----------------------------------
         No |         32       47.76       47.76
        Yes |         30       44.78       92.54
            |          3        4.48       97.01
 Don't know |          2        2.99      100.00
------------+-----------------------------------
      Total |         67      100.00
```

Did you get any federal funds to cover staff/volunteer/participant time/incentives to distribute naloxone (include federal funds passed through the state/county/city)?

In [25]:
```
tab v4, m sort
```

```
   In 2020, |
did you get |
any federal |
   funds to |
      cover |
staff/volun |
teer/partic |
 ipant time |      Freq.     Percent        Cum.
------------+-----------------------------------
         No |         30       44.78       44.78
        Yes |         29       43.28       88.06
 Don't know |          5        7.46       95.52
            |          3        4.48      100.00
------------+-----------------------------------
      Total |         67      100.00
```

"Did you get any state, county, or city government funds to purchase naloxone (exclude federal funds passed through the state, county, or city)?"

In [26]:
```
tab in2020didyougetanystatecountycit, m sort
```

```
   In 2020, |
did you get |
        any |
state/count |
     y/city |
 government |
   funds to |
   purchase |
   naloxone |      Freq.     Percent        Cum.
------------+-----------------------------------
         No |         34       50.75       50.75
        Yes |         29       43.28       94.03
            |          3        4.48       98.51
 Don't know |          1        1.49      100.00
------------+-----------------------------------
      Total |         67      100.00
```

"Did you get any state/county/city government funds to cover staff/volunteer/participant time/incentives to distribute naloxone (exclude federal funds passed through the state/county/city)?"

In [27]:
```
tab v7, m sort
```

```
   In 2020, |
did you get |
        any |
state/count |
     y/city |
 government |
   funds to |
      cover |
staff/volun |      Freq.     Percent        Cum.
------------+-----------------------------------
         No |         34       50.75       50.75
        Yes |         28       41.79       92.54
            |          3        4.48       97.01
 Don't know |          2        2.99      100.00
------------+-----------------------------------
      Total |         67      100.00
```

"Did you have to do fundraising to get enough money OR use unrestricted donations to purchase naloxone?"

In [28]:
```
tab in2020didyouhavetodofundraisingt, m sort
```

```
   In 2020, |
    did you |
  have to do |
fundraising |
     to get |
     enough |
   money OR |
        use |
unrestricte |
          d |      Freq.     Percent        Cum.
------------+-----------------------------------
         No |         31       46.27       46.27
        Yes |         30       44.78       91.04
            |          3        4.48       95.52
 Don't know |          3        4.48      100.00
------------+-----------------------------------
      Total |         67      100.00
```

#### Mutual Aid Redistribution Analysis

In [29]:
```
import delimited "/Users/nabarun/Dropbox/Projects/Buyers Club manuscript/OSNN Master 2021 - 2021 backor
der pool.tsv", delimiter(tab) varnames(1) clear

* Drop excess rows with notes
drop if doesyourorganization==""

 * Create dichotomous indicators
 gen got = regexm(doesyourorganization, "contribute")
 gen need = regexm(doesyourorganization, "need")

tab got
tab need
```

```
(encoding automatically selected: ISO-8859-1)
(20 vars, 109 obs)

(23 observations deleted)



        got |      Freq.     Percent        Cum.
------------+-----------------------------------
          0 |         59       68.60       68.60
          1 |         27       31.40      100.00
------------+-----------------------------------
      Total |         86      100.00


       need |      Freq.     Percent        Cum.
------------+-----------------------------------
          0 |         27       31.40       31.40
          1 |         59       68.60      100.00
------------+-----------------------------------
      Total |         86      100.00
```

##### Expired naloxone

In [30]:
```
gen expiredok=regexm(lower(areyouabletoacceptexpirednaloxon),"yes") if need==1
tab expiredok
```

```
(27 missing values generated)


  expiredok |      Freq.     Percent        Cum.
------------+-----------------------------------
          0 |         24       40.68       40.68
          1 |         35       59.32      100.00
------------+-----------------------------------
      Total |         59      100.00
```

##### Funding

In [31]:
```
tab doyouneedmoneyforshipping doyouneedmoneyforstorage
```

```
    Do you |
need money | Do you need money for
       for |        storage?
 shipping?  |        No        Yes |     Total
-----------+----------------------+----------
        No |        11          0 |        11
       Yes |        14          1 |        15
-----------+----------------------+----------
     Total |        25          1 |        26
```

##### Formulation Preferences

In [32]:
```
gen narcan=regexm(lower(whatkindofnaloxonecanyouacceptch),"narcan") if need==1
    di "Nasal Narcan"
tab narcan
        gen generic=regexm(lower(whatkindofnaloxonecanyouacceptch),"generic") if need==1
    di "Generic injectable"
tab generic
        gen evzio=regexm(lower(whatkindofnaloxonecanyouacceptch),"evzio") if need==1
    di "Evzio autoinjector"
tab evzio
```

```
(27 missing values generated)

Nasal Narcan


     narcan |      Freq.     Percent        Cum.
------------+-----------------------------------
          0 |         17       28.81       28.81
          1 |         42       71.19      100.00
------------+-----------------------------------
      Total |         59      100.00

(27 missing values generated)

Generic injectable


    generic |      Freq.     Percent        Cum.
------------+-----------------------------------
          0 |          3        5.08        5.08
          1 |         56       94.92      100.00
------------+-----------------------------------
      Total |         59      100.00

(27 missing values generated)

Evzio autoinjector


      evzio |      Freq.     Percent        Cum.
------------+-----------------------------------
          0 |         37       62.71       62.71
          1 |         22       37.29      100.00
------------+-----------------------------------
      Total |         59      100.00
```

*fin.*
